# A scoping review of remote and unsupervised digital cognitive assessments in preclinical Alzheimer’s disease

**DOI:** 10.1101/2024.09.25.24314349

**Authors:** Sarah E. Polk, Fredrik Öhman, Jason Hassenstab, Alexandra König, Kathryn V. Papp, Michael Schöll, David Berron

**Author notes:** Corresponding authors: Sarah E. Polk, Clinical Cognitive Neuroscience, German Center for Neurodegenerative Diseases (DZNE), Magdeburg, DE & David Berron, Clinical Cognitive Neuroscience, German Center for Neurodegenerative Diseases (DZNE), Magdeburg, DE. MS and DB contributed equally to this manuscript.

## Abstract

Characterizing subtle cognitive changes in preclinical Alzheimer’s disease (AD) is difficult using traditional neuropsychological assessments. Remote and unsupervised digital assessments can improve scalability, measurement reliability, and ecological validity, enabling the capture of subtle changes. We evaluate such tools for use in preclinical AD, or cognitively unimpaired individuals with abnormal levels of AD pathology. We screened 1,904 reports for studies remotely assessing cognition in preclinical AD samples. Twenty-three tools were identified and their usability, reliability, and validity, including construct and criterion validity based on in-person neuropsychological and Aβ/tau measures, was reported. We present a necessary update to a rapidly evolving field, following our previous review (Öhman et al., 2021) and address open questions of feasibility and reliability of remote testing in older adults. Future applications of such tools are discussed, including longitudinal monitoring of cognition, scalable case finding, and individualized prognostics in both clinical trials and healthcare contexts.

## Introduction

Subtle cognitive changes may already emerge in the preclinical stage of Alzheimer’s disease (AD), that is, in cognitively unimpaired individuals with abnormal levels of biomarkers indicative of AD pathology, such as amyloid-β (Aβ) and tau (clinical stage 2 according to ref. ^1^). Determining whether an individual has abnormal levels of Aβ and/or tau, and thereby establishing a probable etiology for cognitive decline and later impairment, can now reliably be achieved using fluid and imaging biomarkers^1,2^.

However, the characterization of cognitive changes has proven to be challenging using conventional pen-and-paper neuropsychological assessments^3–5^. This presents a non-negligible and potentially costly hurdle, as clinical trials aim to screen and enroll participants at risk for disease-driven cognitive decline before the clinical symptoms of AD appear^6^. Indeed, Langford and colleagues^7^ estimated that using sensitive cognitive measures to pre-screen individuals before ordering PET imaging could have saved over 3.5M USD in the A4 study. Additionally, although there is yet no medical intervention approved for preclinical AD, the use of anti-amyloid drugs in preclinical samples is already being tested in studies such as AHEAD 3-45 (NCT04468659) and TRAILBLAZER-ALZ 3 (NCT05026866). Once early treatments become available, tools that can reliably detect, predict, and monitor cognitive decline due to AD on an individual level will be essential^8^.

Remote and unsupervised digital cognitive assessments have already shown promise as a widely accessible solution that may be sensitive to subtle cognitive changes related to the initial build-up of AD pathology^9^, even before clinical symptoms manifest (i.e., preclinical AD). So far, in the context of preclinical AD, remote and unsupervised digital cognitive assessments have been exclusively used in research, for example, to characterize cross-sectional cognitive differences between healthy and AD samples (e.g., refs. ^10–16)^. Studies have also thus far focused on establishing the feasibility of remote and unsupervised data collection in samples of older adults with varying degrees of cognitive impairment (e.g., refs. ^17–22^). Moving forward, remote and unsupervised digital cognitive assessments have great potential in future research directions as well as health-care use cases for sensitive cognitive assessments, including in the identification of individuals with subtle cognitive changes (e.g., as a pre-screening for biomarker testing), the quantification of risk of later cognitive decline in individuals (e.g., prognosis), and the characterization of change in cognition longitudinally using frequent assessments (e.g., to monitor cognitive decline and/or treatment outcomes).

Digital data collection, whether remote or in-person, offers a number of advantages over pen-and-paper testing, such as improved measurement precision (e.g., reaction time, automatic scoring)^9^. A wide range of existing pen-and-paper assessments have been digitized^23,24^, and a growing number of cognitive tests are being developed specifically for digital administration (e.g., refs. ^25–27^). In addition to tests that capture conventional cognitive constructs (e.g., episodic memory, processing speed), novel metrics quantifying cognitive function have also been developed, for example using speech-based tasks (e.g., refs. ^28,29^) or multi-modal assessments (i.e., assessment of both active and passive markers of cognition simultaneously^30^). Notably, many tools that use cognitive tests that may be considered conventional can also capture novel metrics (e.g., learning curves using an episodic memory task) due to the digital nature of the tests; thus, this is not a mutually exclusive categorization. Finally, passive data collection can be implemented in participants’ natural environments (e.g., using home-monitoring, wearables), increasing the ecological validity of such metrics by allowing the collection of continuous data streams during individuals’ everyday lives^31,32^. See Figure 1 for an overview of the types of digital cognitive assessments.

**Figure 1.**
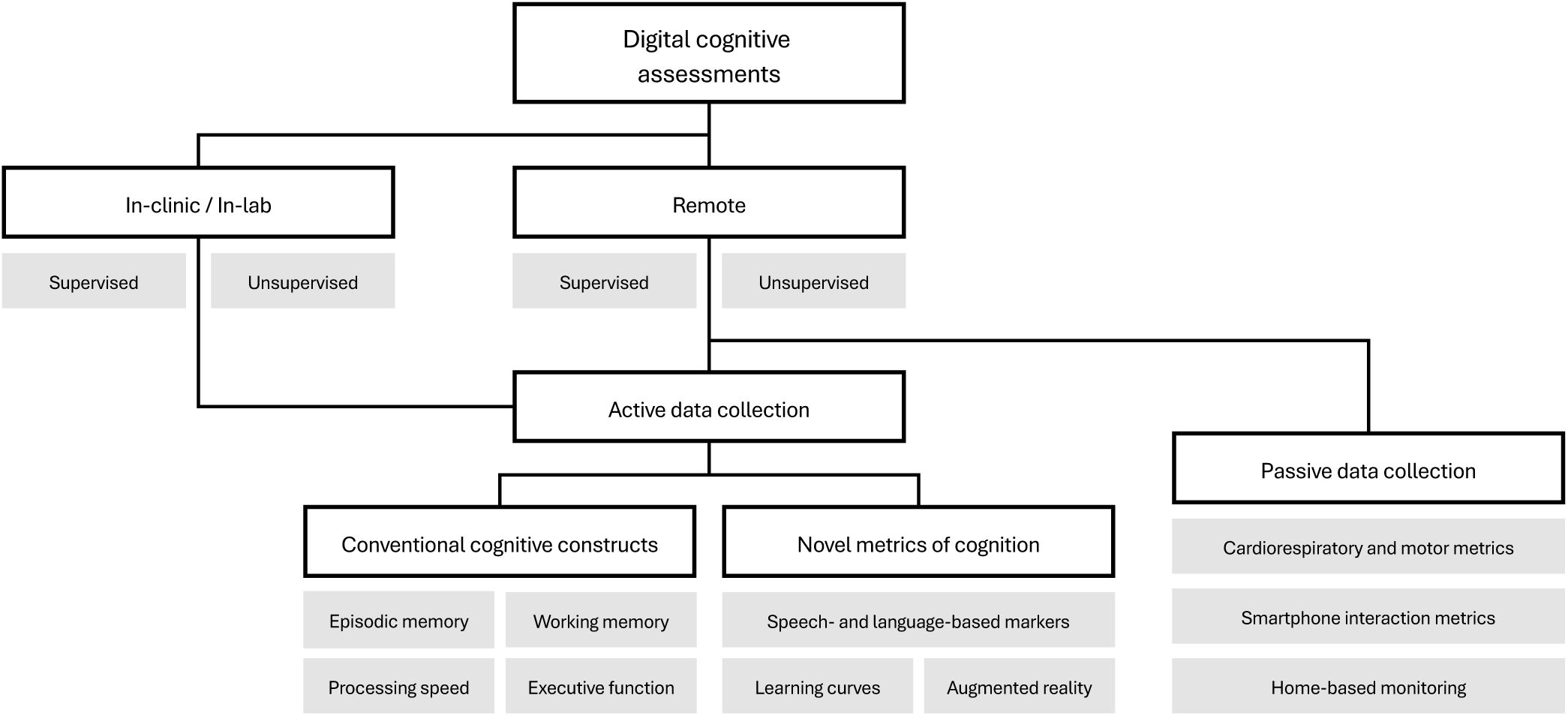
A non-exhaustive taxonomy of the types of digital cognitive assessments. A hierarchical diagram of categories under which digital cognitive assessments may fall in white, with sub-groups and examples of these categories in gray. These categories are not necessarily mutually exclusive. The current review focuses on those assessments that are remotely deployed without supervision, and which use active data collection to quantify cognitive function (i.e., the center of the diagram).

The digitization of cognitive assessments enables the remote and unsupervised collection of cognitive data using participants’ own devices, which is associated with a number of both benefits and challenges. Anyone with a smart device and a network connection can complete remote automated testing. Indeed, data from millions of users across the globe have already been collected using mobile chat- and game-based (e.g., refs. ^27,33^) as well as web-based cognitive tests (e.g., TestMyBrain). This approach offers many benefits, including cost effectiveness and reduction of patient burden while still allowing for a high volume of data to be collected, but also many challenges. For example, remote versus in-person data collection is associated with reduced costs related to travel, clinic or laboratory space, and test administration^34^, though there may be other costs associated with technical support and data servers. Remote data collection can also be less burdensome to patients and participants, as scheduling conflicts (e.g., going into a clinic during business hours) can typically be eliminated and transportation to and from testing sites is not necessary, though participant burden will also depend on the remote testing schedule.

In addition to efficient data collection from more individuals, the frequency of within-person testing can be increased with remote testing. Participants can feasibly self-administer remote assessments once a month for a year^10^, on a daily basis^18,35^, or even multiple times a day^36–38^. Measurement burst designs can also be implemented (e.g., a week of daily assessments every six months for several years)^39,40^. High-frequency testing not only improves the temporal scale on which changes can be detected, but also crucially increases measurement reliability and the sensitivity to intra-individual variability in performance^41–43^. Notably, many remote and unsupervised paradigms that do not explicitly test learning of specific stimuli use randomized stimulus pairs or parallel versions when repeatedly administering tests to reduce retest effects^13,20,37,44–47^. Finally, higher frequency within-person testing has opened the door for novel paradigms measuring cognitive processes that are otherwise difficult to capture, such as learning of repeated stimuli^11,12,35^, recall after several hours or days^46,48,49^, or the effects of time of day^13^.

Another benefit to unsupervised cognitive testing is an increase in ecological validity. One shortcoming of in-clinic tests is potentially discrepant cognitive performance in clinical settings versus at home, known as the “white-coat effect”^50,51^. Performance on remote assessments may be more reflective of individuals’ everyday cognitive function, as it may be less associated with increased anxiety during testing^52^. On the other hand, unproctored testing introduces challenges related to environmental distractions, low effort, and malingering, as well as data fidelity (e.g., ref. ^53^). Building attention checks into tasks and asking explicitly if participants were distracted while completing the tasks may limit these effects. Researchers may also consider implementing algorithms to flag data patterns indicative of cheating, as well as integrating reliable participant authentication processes, data privacy policies permitting.

Another challenge relates to the level of digital literacy necessary for individuals to complete smart device-based testing without assistance, which can vary depending on the target population and tends to be lower in older samples^54^. Additionally, access to a smart device and a dependable network connection may be a limitation, for example, when working with low-income populations or those in remote rural areas.

According to a report published by the International Telecommunication Union^55^, 67% of the global population uses the Internet. Notably, this statistic is highly variable across regions and income levels, and Internet access is particularly limited in Africa compared to other continents, as well as in rural areas and low-income countries^55^.

These statistics should be borne in mind when planning remote data collection and interpreting results, especially considering the strong role that both age and socioeconomic status play in neurodegenerative disease^56,57^.

Finally, the underlying infrastructure of and resources dedicated to data storage and handling will impose an upper limit on the scalability of any remote data collection, and the implementation of big data storage technologies^58^ should be considered sooner rather than later. As such data storage and transfer systems are engineered, particular thought should be given to data privacy and protection, as individual cognitive health data is highly sensitive^59,60^. Researchers should understand the risks as well as costs associated with data transfer and storage to make informed decisions that comply with relevant regulatory frameworks.

With these benefits and challenges in mind, as well as the current need for sensitive cognitive testing in preclinical AD, this scoping review will provide an overview of available remote and unsupervised digital cognitive assessments used in individuals with no clinically identified cognitive decline, but with biomarker evidence for elevated levels of Aβ, or Aβ and tau. We aim to present a necessary update of this rapidly evolving field since the publication of our previous review^9^, with a particular focus on remote and unsupervised assessments rather than digital cognitive assessments as a whole. Open questions in 2021 included whether such remote and unsupervised assessments are feasible in preclinical AD samples, as well as whether such measurements are reliable, especially longitudinally. We discuss progress in the field in this regard, evaluating feasibility based on rates of consent, enrollment, adherence, and compliance, as well as user experience reported by participants (also known as usability validity in the industry-oriented V3+ Framework^61^, which was developed as a common framework for digital health technologies measuring a wide range of metrics; https://datacc.dimesociety.org/v3/), and report the reliability (between- and within-person, parallel-forms) of each tool. We evaluate construct validity (analytical validity in the V3+ Framework) based on associations with established neuropsychological assessments. Regarding the validity for various use cases in preclinical AD (known as clinical validity in the V3+ Framework), we describe whether they can accurately classify individuals with elevated Aβ (and tau) burden, whether they correlate with continuous measures of Aβ and tau, and whether they can predict future cognitive decline. To date, very few studies have used passively collected markers to characterize cognition in preclinical AD^31,32^, therefore we focus on studies using actively collected digital assessments in the current review. Finally, since the feasibility, reliability and validity of remote and unsupervised cognitive testing for use in preclinical AD have been established conceptually, we discuss the direction in which the field is quickly moving, addressing important future use cases, such as scalable case-finding, longitudinal monitoring, and individualized risk assessment, as well as what should be considered in the development of future cognitive tools.

## Results

### Literature screening

A total of 2,688 records were found, 784 of which were excluded as duplicates or records in languages other than English. The remaining 1,904 were screened and 28 relevant reports were found (see Figure 2 for details), two of which were previously included^9^. These and the 23 tools described in these records are listed in Table 1.

**Figure 2.**
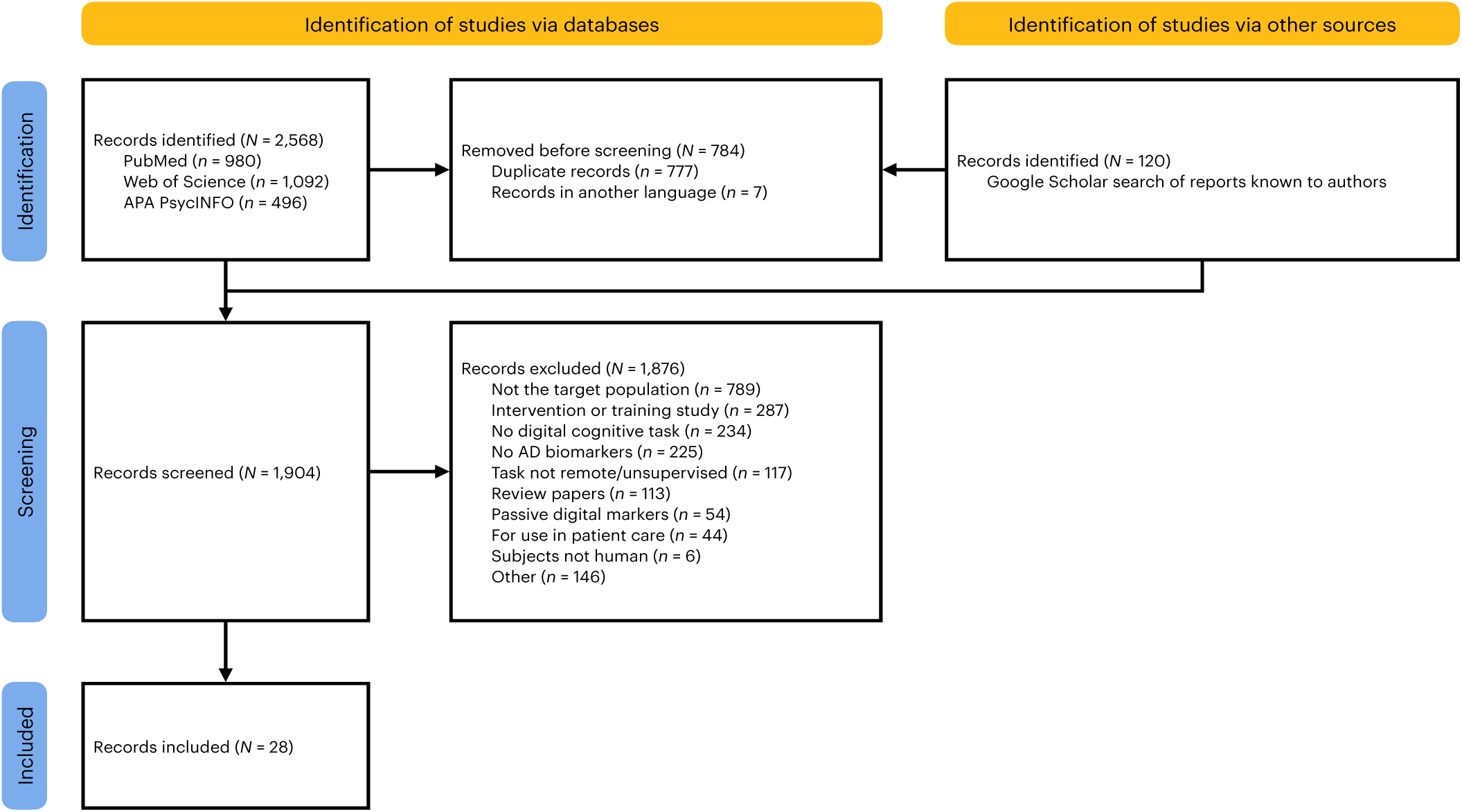
Preferred Reporting Items for Systematic reviews and Meta-Analyses (PRISMA) flow diagram detailing the screening of records. Numbers of records identified from which sources (databases and other), pre-screening of duplicates and records in a language other than English, screening and exclusion of records (with reasons), and inclusion of records are detailed.

**Table 1.**
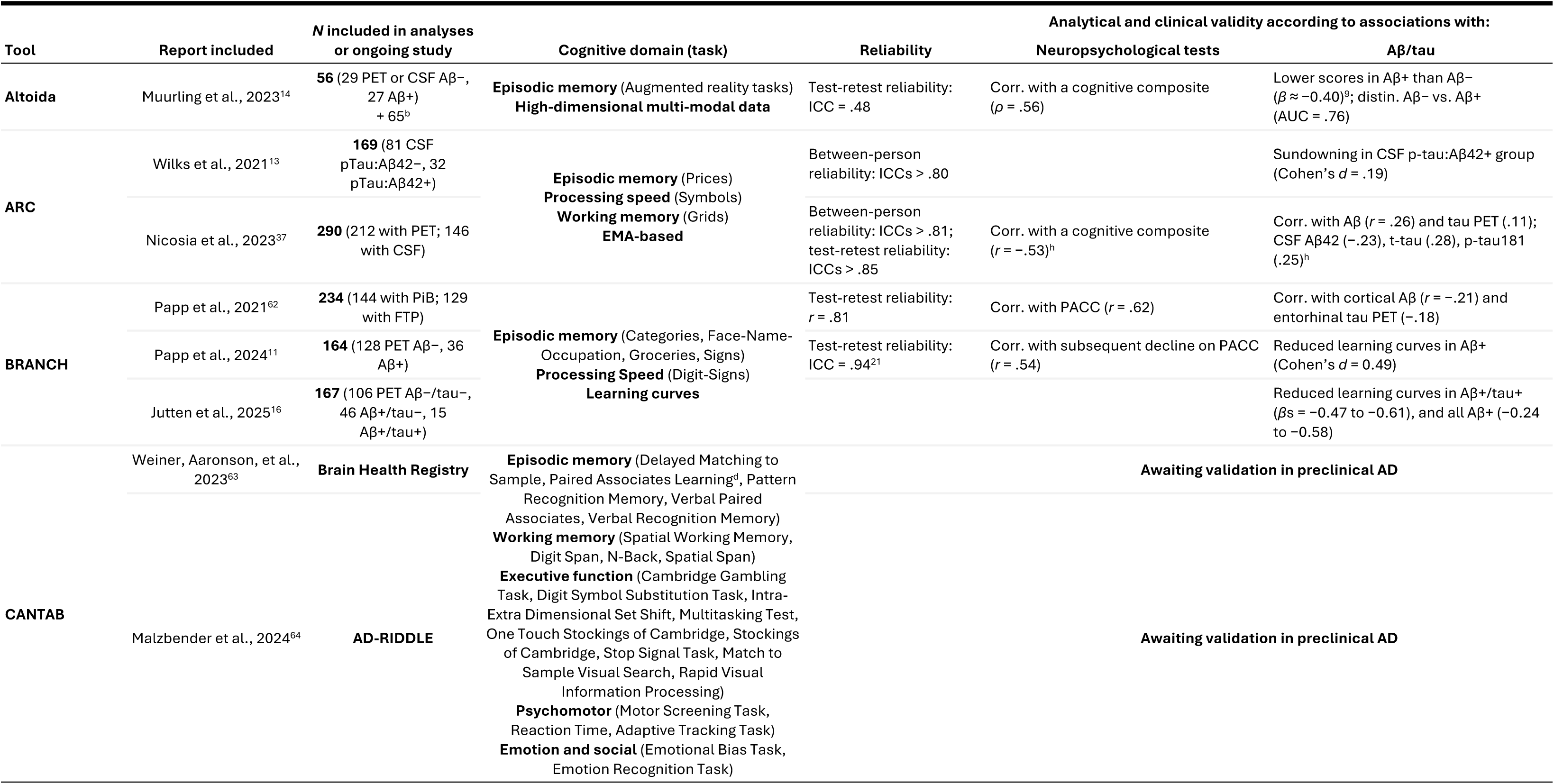
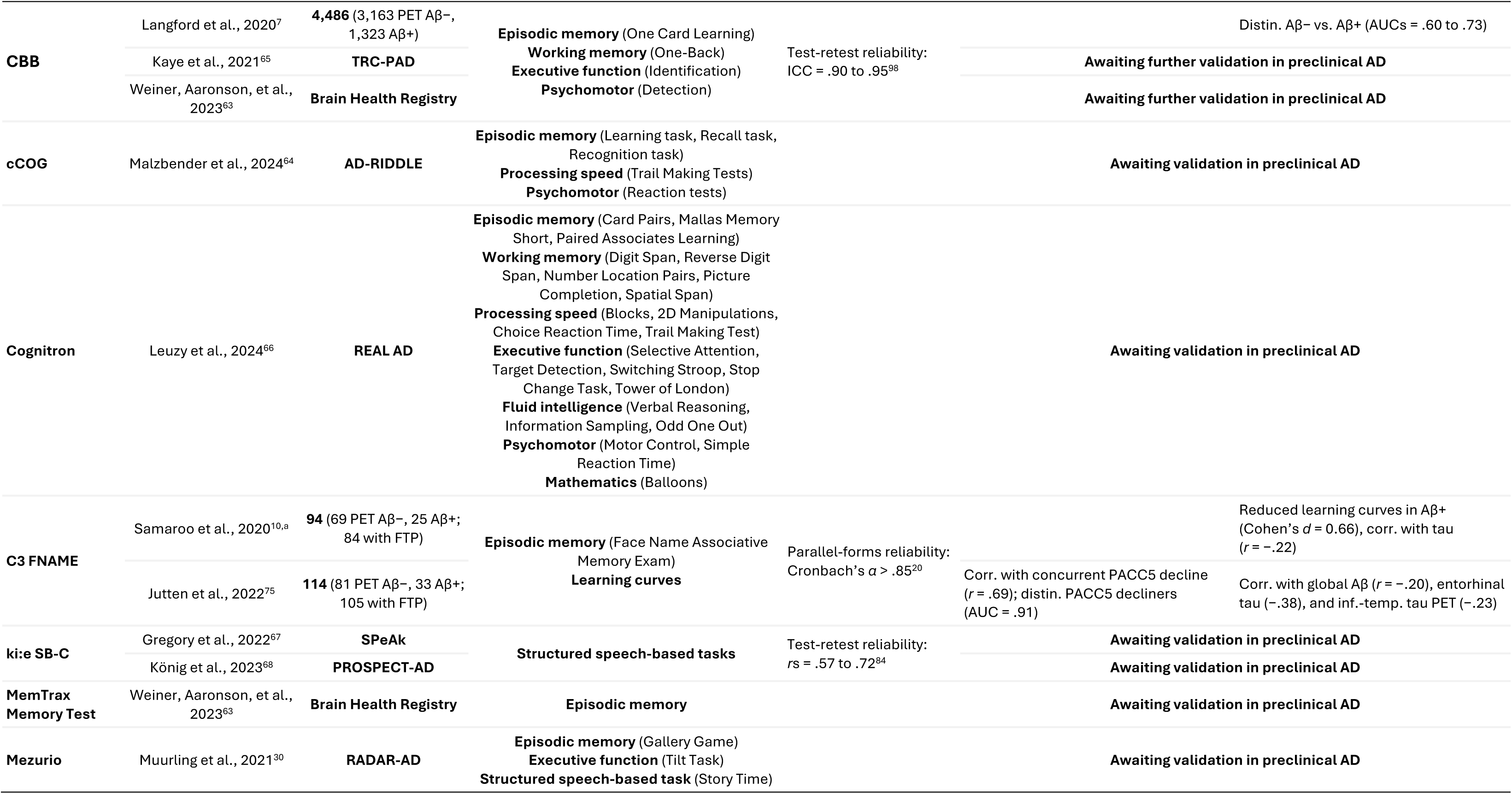
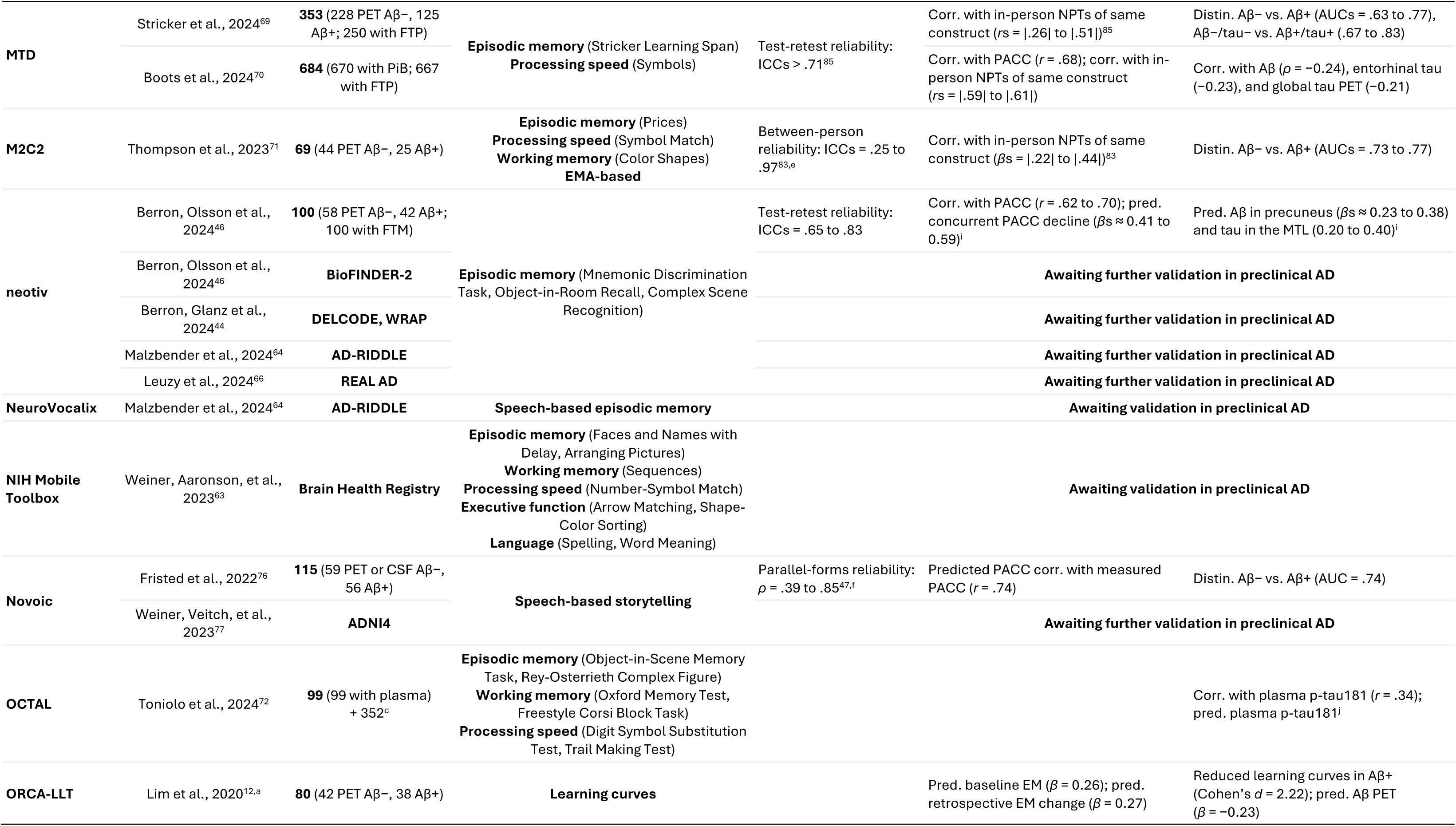
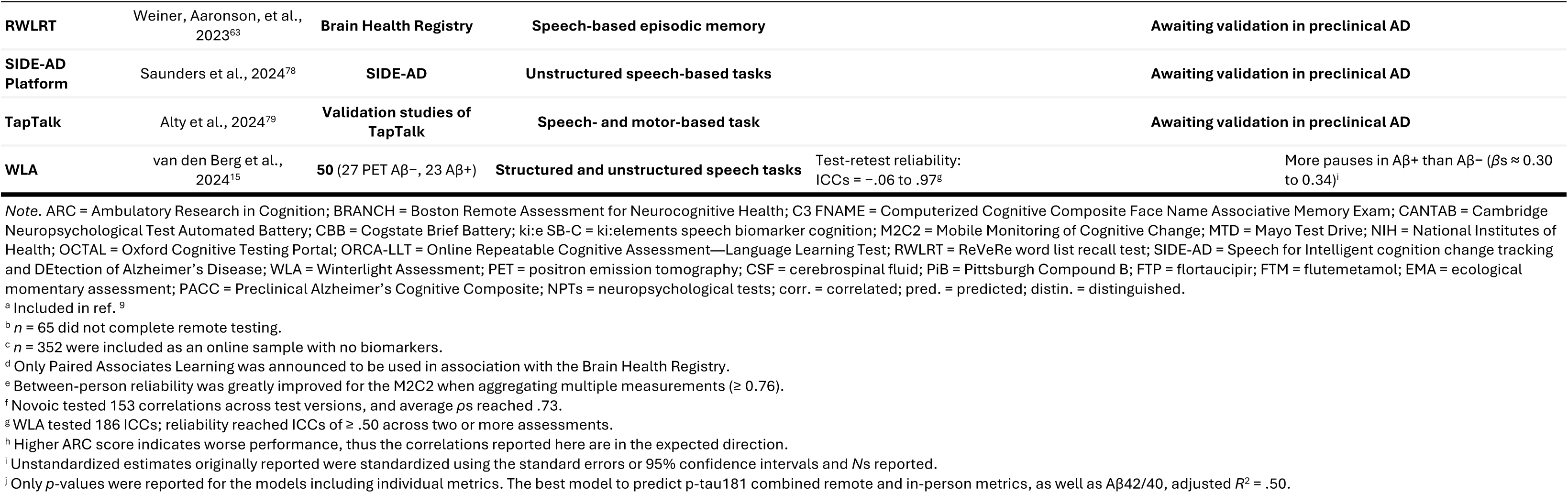
Remote and unsupervised digital cognitive assessments for use in preclinical AD samples in alphabetical order. For samples including groups other than preclinical AD, effect sizes are reported for the findings pertaining to the preclinical AD sub-sample.

### Remote capture of both conventional and novel metrics of cognition

Seventeen of the included tools capture the conventional cognitive constructs: Altoida^14^, Ambulatory Research in Cognition (ARC)^13,37^, Boston Remote Assessment for Neurocognitive Health (BRANCH)^62^, Cambridge Neuropsychological Test Automated Battery (CANTAB)^63,64^, Cogstate Brief Battery (CBB)^7,63,65^, cCOG^64^, Cognitron^66^, the ki:elements speech biomarker for cognition (ki:e SB-C)^67,68^, Mayo Test Drive (MTD)^69,70^, MemTrax Memory Test^63^, Mezurio^30^, Mobile Monitoring of Cognitive Change (M2C2)^71^, neotiv^44,46,64,66^, NeuroVocalix^64^, NIH Mobile Toolbox^63^, Oxford Cognitive Testing Portal (OCTAL)^72^, and the ReVeRe word list recall test (RWLRT)^63^. One of these tools (Altoida) uses an augmented reality-based cognitive task to collect multi-modal data (i.e., simultaneous capture of active and passive markers during a cognitive task), one (BRANCH) is used to quantify learning of repeated stimuli over days to months (i.e., learning curves)^11,16^, four (ki:e SB-C, Mezurio, NeuroVocalix, RWLRT) evaluate cognitive function using speech-based metrics, and two (ARC and M2C2) use ecological momentary assessment (EMA)-based paradigms^73^ (EMA is a method of frequently sampling variables at random times of the day while subjects are in their natural environments to capture intra-individual variability; see refs. ^73,74^). Two other tools are used to quantify learning curves: Computerized Cognitive Composite (C3) Face Name Associative Memory Exam (FNAME)^10,75^, and Online Repeatable Cognitive Assessment—Language Learning Test (ORCA-LLT) ^12^. Finally, an additional four tools use speech-based metrics: Novoic^76,77^, the Speech for Intelligent cognition change tracking and DEtection of Alzheimer’s Disease (SIDE-AD) online platform^78^, Winterlight Assessment (WLA)^15^, and TapTalk^79^, the last of which combines speech and motor function. Nine of these tools were discussed in the previous review either as in-person (e.g., CANTAB, NIH Toolbox, Cogstate) or remote digital tools^9^, but had not yet been validated for remote use in preclinical AD (except BRANCH and ORCA-LLT^10,12^).

### High feasibility of remote and unsupervised data collection

These studies were generally highly feasible, as shown by rates of consent into studies, enrollment, adherence, compliance, and user experience reports. Three studies, all of which recruited from ongoing studies, reported the consent rate of those individuals approached for participation: a year-long learning curves design with monthly assessments had a consent rate of 86%^10^, and a week-long measurement burst design^39,40^ with four daily measurements and had a consent rate of 87%^54^. Finally, a longitudinal paradigm with biweekly assessments for 12 months also reported an 86% consent rate^46^. However, the latter also reported that 24% of those who consented did not enroll in the app^46^.

Adherence, generally measured as the average percentage of tests completed out of the complete study protocol, ranged from 74% to 93% for week-long protocols with up to four daily measurements^11,13,15,37,71,76^, from 63% to 94% for an eight-week study^19^, and from 75% to 78% in year-long studies^12,75^. Compliance, defined here as the completion of measurements as intended (e.g., acceptable data quality, successful attention checks), was generally excellent, with only 2 to 3% of data being unusable in both cross-sectional and longitudinal designs measuring conventional cognitive metrics as well as learning curves^15,62,72,75^. However, 21% of data from the in-home augmented reality tasks were unusable due to technical issues, and another 32% of participants were unable to complete the in-home tasks either because their smartphone was not compatible or for other unspecified reasons^14^. Similarly, preliminary feasibility results from the current Alzheimer’s Disease Neuroimaging Initiative cohort (ADNI4) showed that only 54% of eligible individuals completed speech-based tasks^80^.

Regarding user experience, in a cross-sectional paradigm, 16% of participants reported they had technical difficulties^62^, and in a week-long speech-based paradigm, 15% reported difficulties^47^, while in a year-long design, 30% of participants reached out for technical support^10^. Tools were generally straightforward and enjoyable to use^11,13,15,37,46,47,62,71,81^, with 65% of one sample reporting that they would “definitely” complete such sessions again^38^. Additionally, a majority of participants asked indicated that they preferred remote testing to pen-and-paper testing (75%^46^ and “most”^37^).

### Acceptable reliability of remote and unsupervised data collection

We also evaluated the reliability of each tool, insofar as this was reported (see Table 1 for specific statistics). Between-person reliability (i.e., variability across individuals, calculated with intraclass correlations [ICCs] according to ref. ^82^) was good for ARC and across multiple M2C2 assessments, with ICCs of 0.76 or greater^13,37,83^.

Parallel-forms reliability (i.e., reliability across alternate versions of the same task) was good for C3 FNAME (Cronbach’s *α* > .85)^20^, as well as for Novoic on average across versions (mean *ρ* = .73)^76^; no other studies reported parallel-forms reliability. Test-retest reliability (i.e., precision of measurement within an individual) was moderate to excellent for most tasks (ICCs = .65 to .95, *r*s = .57 to .81)^21,37,46,62,84,85^, poor for Altoida (ICC = .48)^14^, and varied widely across WLA metrics (ICCs = −.06 to .97)^15^, though most reached ICCs ≥ .50 across multiple assessments.

### Construct and criterion validity of remote and unsupervised data collection

Finally, we examined the construct validity of each tool based on associations with in-person neuropsychological tests, as well as criterion validity for use in preclinical AD, based on associations with biomarkers of AD pathology (see Table 1).

A number of tools reported cross-sectional associations with established in-person cognitive measures. Those tools that reported associations with measures of global cognition (e.g., Preclinical Alzheimer’s Cognitive Composite [PACC]^86,87^ or similar composites) found correlations coefficients (*r*s and *ρ*s) between |.53| and |.70| and a standardized *β* estimate of 0.26^14,37,46,62^. Two studies found associations between remote tasks and traditional neuropsychological tests measuring the same cognitive constructs (*r*s = |.59| to |.61|; *β*s = |.22| to |.44|)^83,85^. Another study used speech-based metrics to predict PACC5 and found that predicted and measured PACC5 scores correlated (*r* = .74)^76^. Additionally, some studies looked at the associations between remote task performance (cross-sectional and learning curves) and change in a cognitive composite (PACC), with change being retrospective, concurrent, or subsequent relative to the remote task administration (*r*s = .54 to .69, *β*s = 0.27 to 0.59)^11,12,75^. Finally, one tool was able to distinguish those with an annual PACC score decline greater than 0.10 SD from non-decliners (AUC = .91)^75^.

Regarding associations with biomarkers of Aβ and/or tau pathology, a number of studies reported differences in performance between biomarker-negative and -positive groups^10–16^. Most group differences had medium effect sizes (Cohen’s *d*s = 0.49 to 0.66, Hedge’s *g*s = .43 to .63, *β*s = |0.30| to |0.40|), with Aβ+ (or Aβ+/tau+) groups performing worse than Aβ− groups. One study found a small effect of time of day (Cohen’s *d* = 0.19), with individuals with elevated p-tau181/Aβ42 levels performing worse in the evening^13^, while another found a very large effect of Aβ on learning curves across a year, with a Aβ+ group showing dramatically slowed learning (Cohen’s *d* = 2.22)^12^. A number of studies reported associations between performance on remote tasks and continuous measures of Aβ and/or tau^10,12,37,46,62,70,72,75^, with correlation coefficients ranging from |.11| to |.34| and *β*s ranging from |0.23| to |0.38|. Finally, some studies reported that remote task performance could distinguish between Aβ− and Aβ+ individuals^14,69,71,76^ or Aβ/tau− and Aβ/tau+ individuals^69^, with areas under the curve between .63 and .83.

### Remote and unsupervised tools awaiting validation in preclinical AD

A number of tools included in the current review have yet to be validated for use in preclinical AD, or are undergoing further validation for additional use cases, but their deployment in relevant samples has been announced (see Table 1). These tools are included in moderately sized to large studies collecting longitudinal data (e.g., ADNI4, BioFINDER-2, DELCODE, PROSPECT-AD, RADAR-AD, REAL AD, SIDE-AD, SPeAk, WRAP), in registries that collaborate with such studies (e.g., Brain Health Registry), and in large collaborative efforts to inform the research and treatment of AD (e.g., AD-RIDDLE). They include tools that capture conventional cognitive constructs as well as novel metrics of cognitive function, predominantly speech-based markers. All of these tools are deployed in samples that include biomarker-characterized cognitively healthy older adults, and results regarding the validation for their use in preclinical AD are expected.

## Discussion

In this review, we focused on remote and unsupervised digital cognitive assessment tools that capture both conventional cognitive constructs and novel metrics with active data collection to characterize and detect subtle cognitive decline in preclinical AD (see Figure 3). We defined preclinical AD as the absence of clinically established cognitive impairment in the presence of markers of Aβ, and sometimes tau, pathology. We found that remote and unsupervised cognitive assessments generally have good feasibility and validity for use in preclinical AD, and that the field is quickly moving forward with larger samples and longitudinal studies to address relevant use cases in both clinical trials and health care contexts.

**Figure 3.**
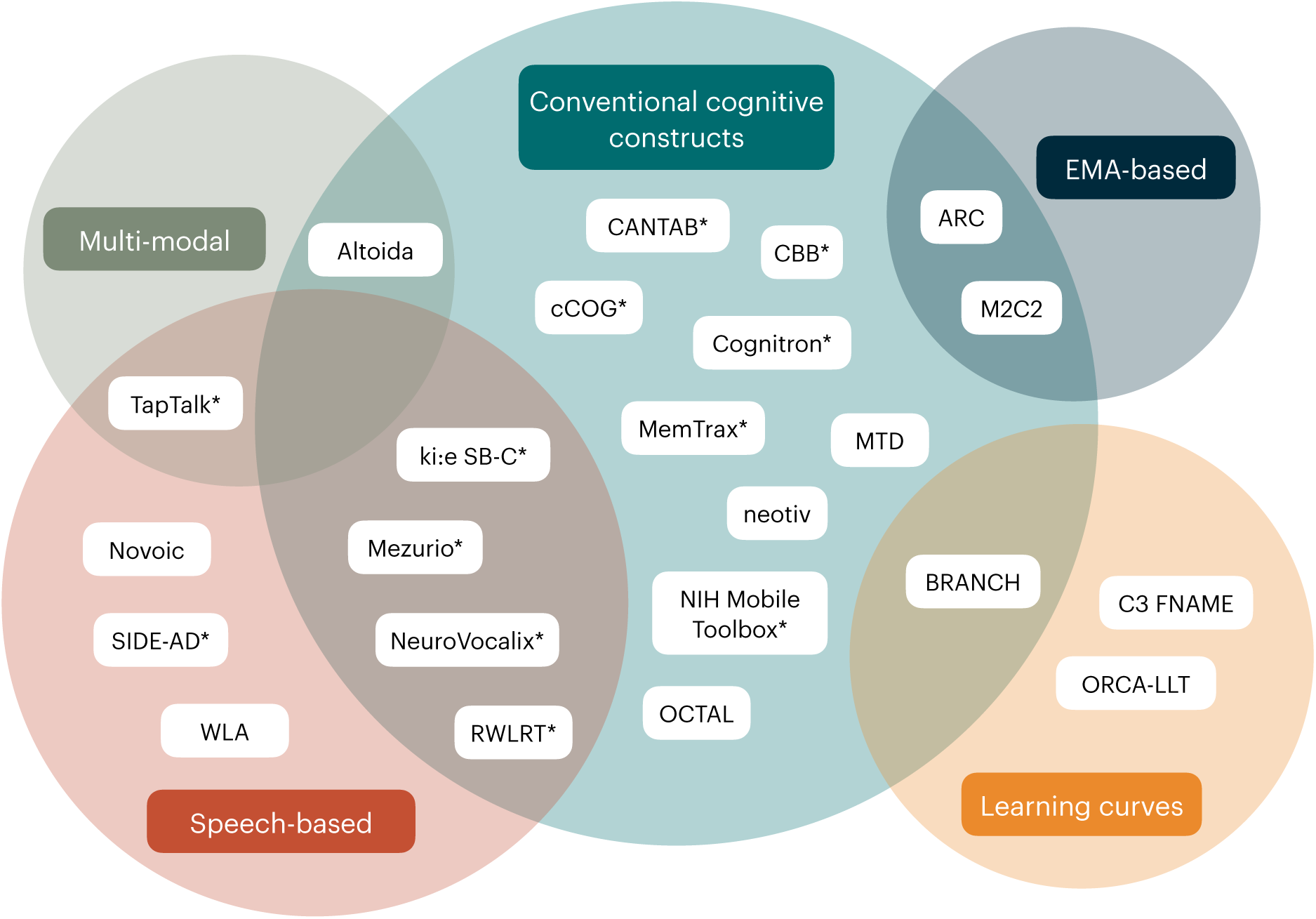
Venn diagram of the tools included in the current review based on the type of cognitive metrics they quantify. Tools identified in the current scoping review were categorized according to their methodology and metrics that they quantified. Tools measuring conventional cognitive constructs are shown in the teal circle; tools using multi-modal data collection are shown in the green circle; tool capturing speech-based metrics are shown in the red circle; tools using EMA-based protocols are shown in the dark blue circle; and tools quantifying learning curves are shown in the yellow circle. Some tools are considered to belong to multiple categories. *EMA = ecological momentary assessment; ARC = Ambulatory Research in Cognition; BRANCH = Boston Remote Assessment for Neurocognitive Health; C3 FNAME = Computerized Cognitive Composite Face Name Associative Memory Exam; CANTAB = Cambridge Neuropsychological Test Automated Battery; CBB = Cogstate Brief Battery; ki:e SB-C = ki:elements speech biomarker cognition; M2C2 = Mobile Monitoring of Cognitive Change; MTD = Mayo Test Drive; NIH = National Institutes of Health; OCTAL = Oxford Cognitive Testing Portal; ORCA-LLT = Online Repeatable Cognitive Assessment—Language Learning Test; RWLRT = ReVeRe word list recall test; SIDE-AD = Speech for Intelligent cognition change tracking and DEtection of Alzheimer’s Disease; WLA = Winterlight Assessment. *Tools awaiting validation for use in preclinical AD samples*.

At the time our previous review was published^9^, the question of whether it is feasible to remotely deploy digital cognitive tools without supervision in preclinical AD samples was still open. Among the studies covered in the current review (two that were previously included^10,12^) we found that rates of consenting into studies (albeit from other ongoing studies), adherence (i.e., how many measurements participants completed), and compliance (i.e., how many measurements participants completed *as intended*) were impressive. Across study designs ranging from one week to one year, consent rates ranged from 86 to 87%, adherence rates from 63 to 93%, and compliance rates from 97 to 98%. This is in stark contrast to a previous report of the median participant retention in digital health studies being only 5.5 days out of 12 weeks^88^.

Another indicator of usability was the generally positive user feedback collected with user experience surveys. However, it is important to note that many digital assessment studies included here recruited from existing longitudinal study cohorts; participants in these studies typically agree to complete fluid biomarker acquisition, neuroimaging, and regular in-clinic assessments and therefore do not represent the general population. Adherence reported in the studies included in this review may therefore be inflated compared to the expected adherence to such paradigms among real-world samples.

One potential bottleneck to participant retention may be registration in digital apps after consenting into a study. For example, out of those who consented to participate in a year-long study, only 64% actually registered in the smartphone app and completed at least one task^46^. Similarly, of those individuals who met ADNI4 inclusion criteria, only 54% completed speech-based remote tasks^80^. One factor that may ameliorate participant drop-out is the technical support available; while 15 to 16% of participants in cross-sectional or week-long studies reported technical difficulties^47,62^, 30% of participants in a year-long longitudinal study reached out to the study team for technical support^10^. Additionally, as the tasks become more technologically demanding, feasibility may become limited, as was the case for the augmented reality-based tasks, during which 21% of participants had technical issues precluding the use of their data, and another 32% could not complete the tasks because of smartphone incompatibility or other unnamed reasons^14^. Indeed, when issues with remote technology were quantified (from 0 to 1, 0 = no problems) for both active cognitive tools included in RADAR-AD, the same participants had fewer technology-based problems with the app used for administering conventional cognitive tasks (.31) than with the augmented reality-based app (.60)^19^.

As more and more large longitudinal studies implement remote data collection, special attention should be paid to participant enrollment and retention, especially when recruitment is done remotely and outside of Western, Educated, Industrial, Rich, and Democratic (WEIRD) populations. One way of boosting retention, for example, is by having participants engage with the digital tool immediately after consenting into a study^89^. Having technical support available to participants, especially those with lower digital literacy^54^, may also alleviate both issues of limited enrollment as well as prohibitive technical difficulties during tasks.

Another open point in the previous review pertained to the reliability of remotely assessed cognitive metrics^9^. The reporting of reliability varied across studies, but reported reliability was generally good. Three studies reported between-person reliability (i.e., the ability of the test to differentiate individuals), two using ARC and one using M2C2^13,37,83^; between-person reliability of ARC was high, as well as across multiple sessions of M2C2. Parallel-forms reliability was only reported by two studies: C3 FNAME showed good reliability across alternate versions^20^, while Novoic tasks had moderate to excellent reliability across versions^47^. Test-retest reliability (i.e., the reliability of a test within an individual across multiple assessments) was more widely reported, with moderate to excellent reliability of five tools capturing conventional cognitive constructs as well as learning curves^21,37,46,62,85^. The test-retest reliability for the speech-based WLA varied depending on the metric and the number of assessments across which performance was averaged^15^. For both neotiv and WLA, test-retest reliability improved when aggregating across multiple measurements^15,46^. Altoida was the only tool that reported poor retest reliability^14^, which was acceptable for Android users, ICC = .70, but very poor for iOS users, ICC = .33. Authors speculated that there may have been version effects, the sample may have been too small to accurately capture test-retest reliability (*n* = 43), or participants may have received help during some measurements but not others^14^.

Overall, reliability was generally favorable, suggesting that cognitive testing can reliably separate individuals and also precisely capture cognition within the same individual across multiple measurements, even when done remotely and without supervision. As longitudinal data collection and the quantification of cognitive changes becomes the goal of more and more studies, reliability of assessment tools, especially test-retest reliability, should be carefully assessed in all target populations, and reasons for poor reliability should be corrected to ensure that the capture of subtle change is not obscured by random noise.

Regarding construct validity, the cross-sectional associations between remotely administered digital cognitive tools and in-person neuropsychological batteries indicate that these remote tools generally map onto cognitive constructs that are important in the clinical quantification of cognitive decline. Additionally, three studies found associations between remote task performance (including learning curves) and change in PACC score, indicating remote task performance may be useful as a prognostic tool to predict cognitive decline. In general, associations were moderate to strong, however no correlations greater than .70 were found. As a benchmark, we looked at digital tests that had been administered remotely as well as in-person: Altoida performance was correlated across at-home and in-clinic administration with a Spearman’s *ρ* of .57 ^14^, as was C3 (composite score across all tasks including FNAME) with an *r* of .70^20^, while a remote version of the neotiv Mnemonic Discrimination Task correlated with a similar in-scanner version with an *r* of .66^46^. This suggests that remote and in-clinic assessments, even when using nearly identical measures, may only be correlated to a limited degree.

Regarding these moderate correlations, Stricker and colleagues^69^ argue that correlating novel measures of cognition with established measures is also not a foolproof method of validation, since the existing measures of cognition, although extensively validated, are themselves imperfect^90^. Many existing neuropsychological tests were developed to quantify cognitive impairment in symptomatic individuals years or even decades before the emergence of reliable *in vivo* biomarkers of AD and neuroimaging^90–92^. The constructs that traditional neuropsychological assessments capture may therefore be 1) insensitive to changes in non-symptomatic individuals (e.g., cognitively healthy individuals may reach ceiling) and 2) outdated in terms of our current understanding of the biological progression of AD. In comparison, the development of novel tests of cognition in AD can be tailored to capture subtle changes as well as be neuroanatomically informed (e.g., neotiv^46^). Additionally, the target population should again be taken into account here; for example, there may be reduced digital familiarity among older adults^54^, further limiting correlations between in-person and remote tests. Focusing on other ways of establishing clinical validity, such as using known groups (e.g., cognitively unimpaired Aβ− and Aβ+ groups^69^) may lead to greater precision in measuring behaviors that are meaningful for preclinical AD.

Necessarily, all tools included in this review were used to find associations between cognition and markers of Aβ and/or tau pathology. Most studies used positron emission tomography (PET)-characterized Aβ and tau burden^10–12,14–16,37,46,62,69,71,75,76^, while some used cerebrospinal fluid (CSF) markers, either as the main markers of interest or as a substitute in the case of missing PET scans^13–15,37,76^. Most studies reported known-group validity, comparing Aβ− to Aβ+ groups and finding poorer performance on remote cognitive tests among cognitively healthy individuals with elevated Aβ burden, showing that many of these assessments are sensitive to the subtle cognitive changes in preclinical AD. Associations between continuous measures of Aβ and/or tau and remotely assessed cognitive were also found, suggesting that cognitive performance as captured by these remote and unsupervised tests progressively worsens as AD pathology accumulates in the brain. Altogether, these findings support the claim that remote and unsupervised cognitive assessments can be used to quantify cognitive functions that are affected by AD pathology, even before clinical symptoms emerge.

So far, one study has looked at the associations between blood plasma markers, specifically p-tau181 and Aβ42/40, and cognitive performance on remote digital tasks^72^, which included both cognitively unimpaired individuals as well as those with clinically characterized AD. They found that plasma p-tau181 correlated moderately with a number of digital cognitive markers, while plasma Aβ42/40 only showed weak correlations (correlation coefficients for the whole sample were not reported in ref. ^72^).

Within cognitively unimpaired individuals, a correlation was found between plasma p- tau181 and the *localization time* metric on a “What is where?” task (i.e., associative memory; *r* = .34), and no correlations with plasma Aβ42/40 were observed^72^. Another study combined performance on a remote task with plasma p-tau217 to predict future decline in PACC score in a mostly cognitively unimpaired sample (*β*s ≈ 0.41 to 0.59)^46^. Notably, compared to a model including only demographic factors and plasma p- tau217, a model also including digital cognitive markers explained 7% more variance, with a significant improvement in model fit (difference in Akaike information criteria = −26.430)^46^. These initial findings suggest that blood plasma markers, particularly p-tau markers, and digital cognitive markers may be used in conjunction as scalable and accessible tools to screen for AD in its earliest stages, reducing the need for expensive and invasive procedures such as PET scans, lumbar punctures, and extensive in-person cognitive batteries for individuals with minimal cognitive complaints.

Moving forward, many remote and unsupervised tools are currently undergoing validation for use in longitudinal contexts related to preclinical AD, some with the explicit goal of recruiting more diverse samples (e.g., ADNI4). Once this is achieved, the monitoring of cognitive decline for other use cases, such as safety monitoring and neurocognitive endpoints in clinical trials^8^, will be greatly facilitated. These efforts are especially timely given the recent update to the US Food and Drug Administration’s guidelines regarding drug trials in preclinical AD, in which cognitive endpoints may be sufficient in trials including participants with no detectable cognitive or functional impairment^6^.

Other use cases where remote and unsupervised cognitive assessment may play an integral role in the future include the detection of meaningful cognitive changes within individuals, with the goals of at-scale case finding for both clinical trials (e.g., inclusion into studies) as well as health care contexts (e.g., diagnostic support), and individualized prognoses (i.e., who is at risk for cognitive decline in the future), in addition to the longitudinal monitoring of cognition already discussed.

The use of remote tools for case finding has already been established in small samples. Four studies included here showed that cognitive function captured by remote tools could distinguish between cognitively healthy individuals with and without elevated Aβ burden: two tasks that used conventional cognitive tests, the MTD Stricker Learning Span (verbal learning, which also distinguished between Aβ/tau− and Aβ/tau+)^69^ and the M2C2 tasks (the associative memory task performed best)^71^, Altoida, which used an augmented reality-based task^14^, and Novoic, which employed speech-based tasks^76^. The best performing task achieved an area under the curve of .77 (.83 for Aβ/tau− vs. Aβ/tau+), and while this may not be considered sensitive enough for use as a stand-alone diagnostic instrument, it may be sufficient for use in screening for further confirmatory diagnostic testing, for example, with CSF or PET, or even blood-based biomarkers. Indeed, screening and recruitment for ADNI4 is already facilitated by the use of remote digital cognitive assessments^77^, showing their value as a screening tool for inclusion into AD research.

The abovementioned longitudinal studies will also be a valuable resource in validating the use of remote and unsupervised cognitive assessments for individualized prognostics. Three studies included in this review reported associations between cross-sectional performance on remote tasks or learning curves and concurrent and/or subsequent change in traditional neuropsychological test scores on the group level: performance on the neotiv Mnemonic Discrimination Task and Object-in-Room Recall, especially in combination with plasma p-tau217 and demographic factors, predicted PACC5 decline over up to five years^46^, while learning curves captured with C3 FNAME and BRANCH showed associations with PACC change over one year^11,75^. This suggests that remote cognitive assessments, perhaps in combination with less invasive blood-based biomarkers, may be informative for prognoses in terms of expected future cognitive decline in AD. Future research may seek to validate remote cognitive assessments in terms of how well they can discriminate between individuals who will decline in the future and those whose cognition will remain relatively stable (i.e., individual risk assessment).

The clinical validation of remote and unsupervised cognitive assessment tools for individualized prognostics would be a meaningful milestone for patients and caregivers in particular. The expected onset of clinical and functional decline or the average time until noticeable cognitive symptoms typically emerge or until independent living may become difficult can be valuable information for those affected by neurodegenerative diseases^93^.

Regarding the practical use of current tools, as well as the development of future tools, we found a total 23 tools that are used for remote and unsupervised data collection in preclinical AD, some of which measured overlapping constructs.

Researchers looking to develop new tools to detect early changes in cognition due to AD pathology should review existing tools and their validated use cases to determine whether the development of a new tool is necessary, given the associated monetary costs as well as researcher and participant/patient burden. As a reference for existing digital health technologies used for cognitive assessment as well as other clinically meaningful outcomes and predictors, such as sleep and physical activity, the Digital Health Measurement Collaborative Community (DATAcc) by the Digital Medicine Society (DiMe) has compiled a list of validated digital health tools in the Library of Digital Measurement Products^94^ (https://datacc.dimesociety.org/digital-measurement-library/). As of January 20^th^, 2025, all the tools in Table 1 except OCTAL and ORCA-LLT can be found in the library, and new digital tools are added regularly. The question of which tool is the “best” will depend heavily on the research question and/or use case. While a head-to-head comparison of existing tools is outside of the scope of the current review, it is an important goal to empirically establish which specific tools researchers and clinicians should use, depending on the respective use case, and indeed studies in this direction are already underway (e.g., ref. ^64^).

Any novel tools, especially those developing new markers of cognitive function, should also be subject to rigorous testing of feasibility, reliability, and validity (e.g., following established psychometric procedures and/or the V3+ Framework^61^). In general, tools for the remote and unsupervised assessment of cognitive function should have high compliance and usability, especially within the target population.

They should reliably measure cognitive function across multiple administrations within the same person and, for use cases requiring longitudinal data collection, be relatively resilient to retest effects (unless they specifically aim to quantify practice effects). And finally, they should show associations, cross-sectional and/or longitudinal depending on the use case, with established measures of the same construct, as well as relevant validity for the use case of interest (e.g., case-finding, prognosis, monitoring).

The current scoping review has some limitations that should be acknowledged. Due to the large volume of records found upon the first search, the search process was not continued iteratively^95^. This may have resulted in missed reports, though given the large amount of records screen and the nascency of the field, this is unlikely.

Additionally, only reports in English were included, excluding reports in French, Chinese, German, Portuguese, and Spanish. Future reviews may consider screening reports in all languages for better inclusion of international findings.

In summary, this scoping review identified 28 papers reporting on 23 digital tools for the remote and unsupervised assessment of cognition in preclinical AD. We provided updates to open questions posed by Öhman and colleagues^9^, determining that remote studies of cognition in healthy older adults are largely feasible, with certain restrictions to usability, and that the data collected with such tools are generally reliable, opening the door for the use of such tools longitudinally. Finally, validity has been conceptually established for these tools with respect to their use in preclinical AD and should continue to be evaluated as these tools are implemented in new contexts of use. Currently, studies deploying remote cognitive assessment tools are focused on acquiring larger, more diverse samples over longer periods of time to validate the use of such tools for longitudinal monitoring of cognition. Future goals include exploring how remote and unsupervised digital tools can be used for case-finding on a scalable level—efforts in this regard are already being made (e.g., ADNI4), individualized prognostics and risk assessment—especially as it pertains to those affected by AD, and longitudinal monitoring of subtle changes in the earliest stages of AD.

## Methods

### Search strategy and selection criteria

An initial literature search was performed on September 12^th^, 2023, using PubMed, Web of Science, and APA PsycINFO using terms including digital, remote, unsupervised, smartphone, cognition, Alzheimer’s, and dementia (see Supplementary Methods for exact search terms) with no limitation according to date. Publications already known to the authors were also included. Literature searches were repeated on March 8^th^, 2024, September 9^th^, 2024, and January 14^th^, 2025. All records were uploaded onto Rayyan, a web-based literature screening tool^96^. Duplicates were algorithmically identified, then manually checked and excluded, as were records in languages other than English. The remaining titles and abstracts were screened by SEP and FÖ; ambiguously relevant reports were flagged and reviewed by both authors. For each paper that was excluded, primary reasons for exclusion were recorded.

Peer-reviewed research articles and planned study reports as well as preprints were included if they used self-administered and remote assessments (e.g., no supervision via phone or video call) and if they reported findings related to preclinical AD (i.e., included measures of Aβ and/or tau pathology in cognitively healthy older adults). We included active cognitive tests (i.e., actively completed by participants), excluding those that only measured passive biomarkers (e.g., gait, keystrokes). Studies including other diseases (e.g., cancers, cardiovascular or other neurodegenerative diseases, psychiatric conditions) were excluded, as were studies using the cognitive assessment tool as an intervention.

### Data extraction

Author(s), year of publication, sample size and group allocation (if applicable), name of cohort or ongoing study, names and types of cognitive tasks available, adherence and compliance metrics, usability metrics, reliability metrics were extracted from each of the selected papers. If certain feasibility or psychometric information about the tools was not reported in the included articles, this was gathered from other papers using the same tools, sometimes in separate samples, found using Google Scholar. Associations with in-person neuropsychiatric assessments and measures of AD biomarkers were also charted. Additionally, if unstandardized estimates were reported in the original papers, these estimates were standardized using standard errors or 95% confidence intervals and *N*s to be able to compare effect sizes; these calculations were not done with the original data and are considered approximations.

This manuscript was prepared according to the Preferred Reporting Items for Systematic reviews and Meta-Analyses extension for Scoping Reviews (PRISMA-ScR) Checklist^97^, which can be found in the Supplementary Information.

## Declaration statements

### Data availability

Literature search results can be made available upon request.

### Code availability

Not applicable.

## Acknowledgements

We thank Merle Hinz and Sarah Kriener for their help checking duplicate reports and labeling excluded reports. This work was supported by the NIH/NIA (1R01AG084017-01A1). It is a part of the EU Joint Programme – Neurodegenerative Disease Research (JPND) expert group Remote Digital Assessment and Monitoring for Early Alzheimer’s Disease (REMOTE-AD), which is supported by the German Federal Ministry of Education and Research (BMBF; 01ED2401) under the aegis of JPND (www.jpnd.eu).

## Author contributions

SEP: Literature search, report screening, writing; FÖ: Literature search, report screening, writing; JH: Writing; AK: Writing; KVP: Writing; MS: Conceptualization, writing; DB: Conceptualization, writing. All authors have read and approved the manuscript.

## Supplementary Information

### Supplementary Methods

#### Search terms

((digital*) AND ((remote) OR (unsupervised) OR (ambulatory) OR (self-administered) OR (computer*) OR (smartphone) OR (tablet) OR (iPad) OR (mobile device))) AND (((test*) OR (scor*) OR (assess*) OR (scale*)) AND ((cognition) OR (cognitive) OR (memory))) AND ((older adults) OR (aging) OR (Alzheimer’s) OR (dementia))

**Supplementary Table 1.**
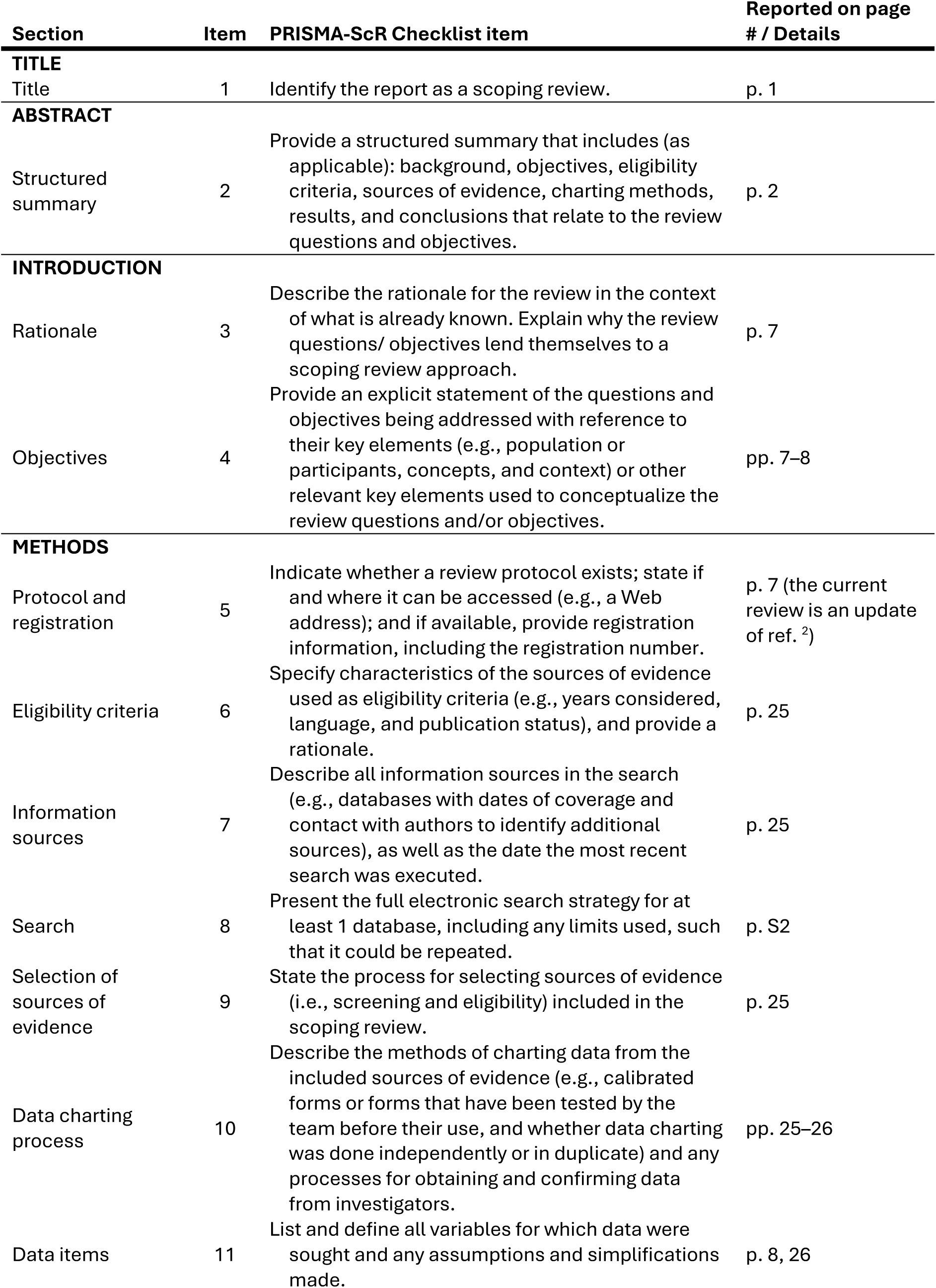
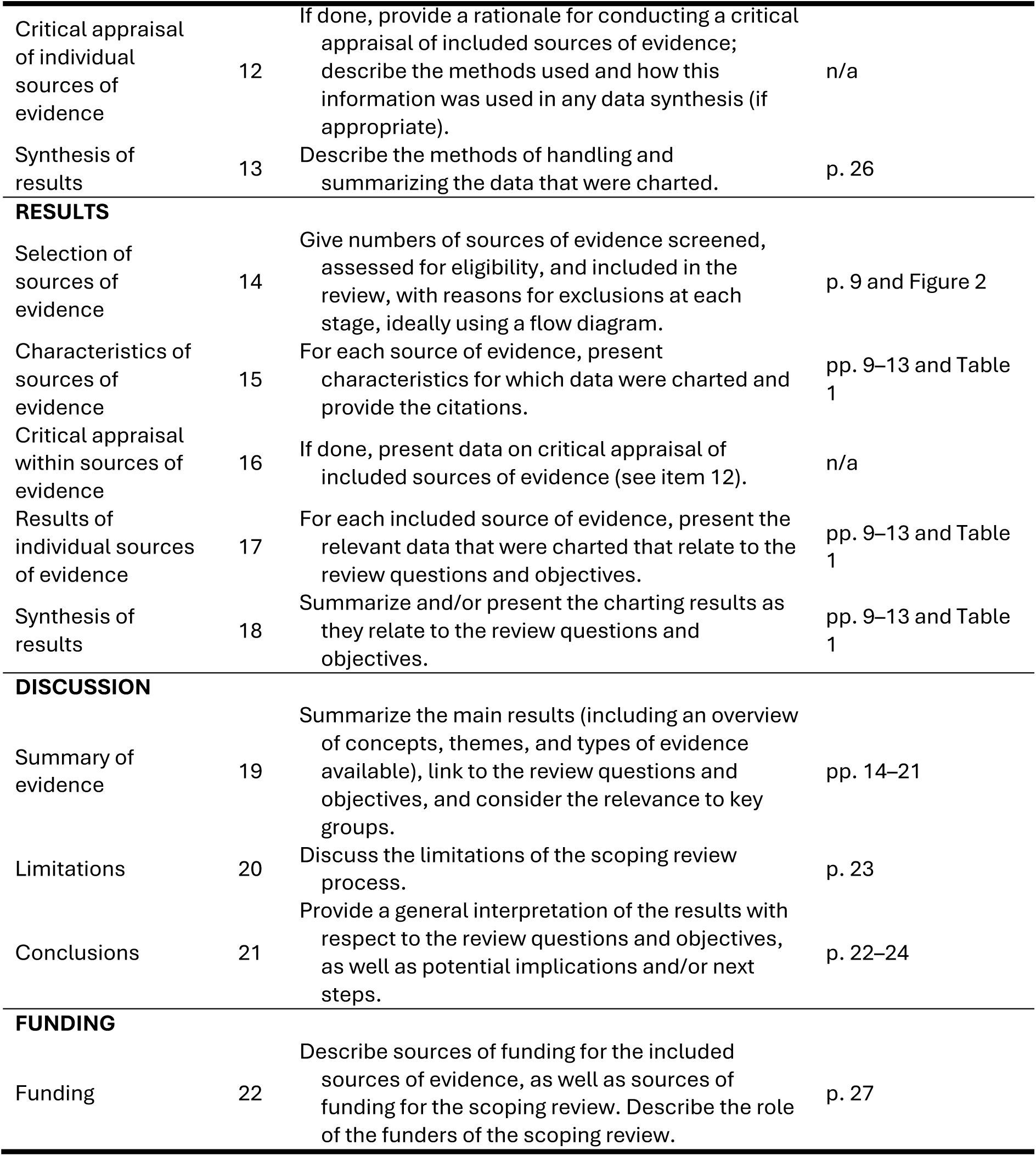
Preferred Reporting Items for Systematic reviews and Meta-Analyses extension for Scoping Reviews (PRISMA-ScR) Checklist according to ref. ^1^.

